# Workplace sexual harassment in sexual and reproductive services: a preliminary cross-sectional study

**DOI:** 10.1101/2021.02.08.21251375

**Authors:** Italo Costanzo

## Abstract

**INTRODUCTION:** Sexual harassment at the workplace in health care is most prevalent in Anglo regions, although is an emerging problem globally. No research has yet focused on the prevalence of the phenomenon within the area of sexual and reproductive health care in Anglo regions. The aim of this study is therefore to measure the prevalence of sexual harassment in sexual and reproductive health care setting and compare it with other clinical areas.

**METHOD:** A web survey to health care workers of various NHS Trusts in the United Kingdom was carried out and a cross-sectional study was conducted to measure observed counts, expected frequencies and prevalence from a total of 90 questionnaires received.

**RESULTS:** The prevalence of workplace sexual harassment within the sexual and reproductive health services is lower compared to other clinical areas.

**CONCLUSIONS:** Working in sexual and reproductive services could be a protective factor for workplace sexual harassment, therefore this study originates a new line of research aiming to identify the protective factors against sexual harassment at the workplace in sexual and reproductive health and the ways they could be used to protect every health care worker from sexual harassment in the workplace.

**KEY POINTS:** The prevalence of workplace sexual harassment in sexual and reproductive health services is lower compared to other clinical areas.

Working in sexual and reproductive services could be a protective factor for workplace sexual harassment.

## BACKGROUND

Any “unwanted, non-reciprocal and unwelcome sexual behaviour that is offensive to those involved and could make people feel threatened, humiliated or even simply embarrassed” is considered sexual harassment[1].

The seriousness of sexual harassment of health care workers has been increasingly the focus of attention and recognised worldwide[2-5].

Although it is estimated that about one in four nurses report having been exposed to sexual harassment in their working life at least once and 17.9% in the previous year[6], the prevalence of sexual harassment against female nurses is 43.15%[7]. Women, who represent the 70% of the health and social workforce[8] are more likely to be sexually harassed in the workplace by people they care for, the family or relatives of patients, and even their co-workers[9-11]; this gender disadvantage is often tolerated for necessity to keep a job by not even reporting incidents[10, 5].

Despite the extreme variability of the prevalence related to the different geographical areas, work environments, research methodologies used, the economic and educational levels and cultural factors[12-13], the total prevalence of workplace sexual harassment in the nursing career is 53.4%[14] with the highest (39%) recorded in Anglo regions, probably because such behaviour appears more socially accepted in places where promiscuous sexual behaviour or outside couple relationship is more accepted[6].

Although exposure to violence appears to be widespread in every healthcare sector[15-16] and differences among various care settings have been found[17], no research has ever focused on sexual harassment at the workplace in the context of sexual and reproductive health care. Addressing sexual health needs of people requires appropriate skills and abilities to be developed so that professionals can easily initiate discussions about the use of contraception, risky behaviours for pregnancy, sexually transmitted infections and HIV[18], as well as reflecting on their beliefs, attitudes and practices in order to be able to understand and recognize them. For its nature, many people could think that working in sexual and reproductive health may involve a higher risk of receiving sexual harassment at work.

Workplace sexual harassment can adversely affect workers ‘psychophysical well-being[19], their motivation, safety, satisfaction, productivity and the efficiency of the whole health organization[20]. It can also impact on patients’ safety, since any aspect related to the assistance, care or treatment necessary for the patient might be omitted, delayed or left unfinished because of the sexual harassment[21-22].

The tendency of not reporting incidents[23-24] reveals the failure of the response of leadership structures in health care organizations[25], as well as the widespread social inability to deal with sexuality[26], despite being largely demonstrated that confrontation with the perpetrator would reduce their disturbing behaviour[27-28] and reporting the incident of violence would help create a safer workplace for everybody and making the work environment free from violence can only be achieved if all the parties involved decide to actively contribute to the changes[25] and without thinking that harassment is an integral part of the job.

The main objectives/aims of this study were the measurement of the prevalence of workplace sexual harassment within sexual and reproductive health care and the statistical comparison with other clinical areas by carrying out a Chi-square independence test. The hypothesis is that working in sexual and reproductive health, dealing daily with different aspects of human sexuality and sexual behaviours, could result in a statistically different risk of sexual harassment at the workplace.

## METHODS

The only eligibility criteria used for this cross-sectional study was being a healthcare worker at the time of filling the survey and it was auto declared by all the participants.

The sample size calculation considered that the authorities enquired (Nursing and Midwifery Council, Office for National Statistics and NHS Digital) had no collected/recorded data on the number of sexual and reproductive healthcare workers in the UK.

The survey was inspired by the questionnaire “Violence in the workplace in the health sector, country case studies, research tools, survey questionnaire”[1], created with a professional software and administered online from August 2020 to September 2020 to the employees of various NHS Trusts in the United Kingdom.

Every individual participated on a voluntary basis, anonymously and gave their implicit informed consent by answering the questions and submitting the survey. An approval from the ethics committee of Cambridgeshire Community Services NHS Trusts was requested and obtained which did not rise any impediment or violation. Confidentiality of all the data processed was guaranteed by anonymity and privacy and an information message was made available at the beginning of the survey, illustrating and explaining purpose and methods of the proposed survey.

## PATIENT AND PUBLIC INVOLVEMENT

No patient involved.

## RESULTS

The number of responses received was 90 and no missed data were found.

The prevalence of sexual harassment at the workplace calculated for the whole population (n = 90) was 28.89%, in the sexual and reproductive health care workers population was 23.08% and in the sample of workers from other clinical areas was 29.87%.

The Chi-squared test falsified the null hypothesis H0 and confirmed that there is a statistically significant difference between the two populations of healthcare workers in sexual and reproductive health services and healthcare workers in other clinical areas.

The odds ratio shows that the risk of sexual harassment at the workplace seems to be lower in sexual and reproductive health services compared to the other areas.

| Working in sexual and reproductive services |  | Experience of sexual harassment |  |  |
| --- | --- | --- | --- | --- |
|  |  | No | Yes | Total |
| No | Observed count | 54.000 | 23.000 | 77.000 |
|  | Expected count | 54.756 | 22.244 | 77.000 |
|  | % within row | 70.130 % | 29.870 % | 100.000 % |
|  | % within column | 84.375 % | 88.462 % | 85.556 % |
|  | % of total | 60.000 % | 25.556 % | 85.556 % |
| Yes | Observed count | 10.000 | 3.000 | 13.000 |
|  | Expected count | 9.244 | 3.756 | 13.000 |
|  | % within row | 76.923 % | 23.077 % | 100.000 % |
|  | % within column | 15.625 % | 11.538 % | 14.444 % |
|  | % of total | 11.111 % | 3.333 % | 14.444 % |
| Total | Observed count | 64.000 | 26.000 | 90.000 |
|  | Expected count | 64.000 | 26.000 | 90.000 |
|  | % within row | 71.111 % | 28.889 % | 100.000 % |
|  | % within column | 100.000 % | 100.000 % | 100.000 % |
|  | % of total | 71.111 % | 28.889 % | 100.000 % |

Contingency Table
|  | Value | df | p |
| --- | --- | --- | --- |
| $X^2$ | 0.250 | 1 | 0.617 |
| N | 90 |  |  |

Chi-Squared Test
|  | Log Odds Ratio | 95% Confidence Intervals |  | p-value |
| --- | --- | --- | --- | --- |
|  |  | Lower | Upper |  |
| Odds ratio | -0.350 | -1.730 | 1.029 |  |
| Fisher's exact test | -0.347 | -2.167 | 1.131 | 0.749 |

## DISCUSSION

Despite the main limit of the study concerning the recruitment of a small number of participants, the study sheds light and offers important data on workplace sexual harassment within the sexual and reproductive services.

Although these findings highlight a whole new aspect of the phenomenon of sexual harassment at the workplace for health care workers and support the idea that sexual harassment can be avoided through educational and training programs that give workers the tools and skills necessary to prevent but also effectively manage the problem[29], the need for further evidence-based research to find new preventive strategies for sexual harassment at the workplace is obvious.

Some of the potential subsequent developments of this study could be the possibilities of investigating on what exactly the specific variables capable of varying the level of risk are, understanding whether the sexual and reproductive health specific knowledge and skills can also be included among these factors and planning to spread these newly founded protective factors to every healthcare worker through appropriate training.

## CONCLUSIONS

The conclusion results in a preliminary evidence that working within sexual and reproductive health services might be a protective factor against sexual harassment at the workplace and suggests how much specialistic knowledge, personal characteristics, skills, aptitude and experience of healthcare workers within sexual and reproductive health services could be a protective factor against the risk of workplace sexual harassment, therefore more research is needed to identify the specific protective variables which could be used to protect every healthcare worker from sexual harassment at the workplace in the future.

## Data Availability

No extra data available.

## ACKNOWLEDGMENTS

Poppy Reynolds (Clinical Nurse Manager, Integrated Contraception & Sexual Health Services, Peterborough), Dr. Paula Waddingham (Senior Research Fellow, Cambridgeshire Community Services NHS Trust), Mike Passfield (Head of Clinical Service, ICaSH), MariaCristina Magnocavallo (Head of Nursing Council of Molise).

